# Clinical characteristics of 2025 Chikungunya outbreak in Chattogram, Bangladesh

**DOI:** 10.1101/2025.09.23.25336475

**Authors:** Mohammed Nasir Uddin, Sanjoy Kanti Biswas, M Jalal Uddin, Taslima Khan Lima, Rajat Sanker Roy Biswas

## Abstract

**Introduction and Objectives:** Chikungunya fever became as a major outbreak in the year 2025 in Chattogram, Bangladesh. Objectives of the present study is to document the clinical profile of the Chikungunya affected patients in Chattogram, Bangladesh.

**Methods:** This is a cross sectional observational study conducted in a period of three months from June 2025 through August 2025 among the clinical, serologically and/or virologically confirmed patients of Chikungunya.

**Results:** Among the 120 patients female was more then the male (52.5% vs 47.5%). Mean age of the patients were found 45.34± 15. 37 years. Most common presentations were fever(81.66%), multiple joint pain (73.33%), ankle swelling and pain (35.83%), bodyaches (43.33%) and rashes (34.16%). Skin rash, nausea/vomiting, urticaria like skin lesions, nose pigmentations, ulcers, morning stiffness were found some as uncommon findings. Haematological study revealed Hb% was normal in 95% patients, leucopenia, neutropenia and lymphopenia were found in 18.33%, 10.83% and 57.50% respectively. Among all, 115(95.83%) had normal ranged platelet. Regarding serological findings Chikungunya IgM and IgG was found positive among 55.0% and 5.83% cases respectively. Regarding molecular data 42(35.0%) were found RT PCR for Chikungunya positive and 5 cases were found for both Chikungunya and Dengue positive. Among the study patients diabetes mellites, hypertension, Ischemic heart disease were found common comorbidities which were 14.1%, 13.3% and 7.5% respectively. One patient was found with RA which made dilemma with post chikungunya arthritis

**Conclusion:** Chikungunya fever showed varied clinical features, serological and molecular findings in 2025 outbreak in Chattogram, Bangladesh.

## Introduction

An arthropod-borne disease Chikungunya which is caused by Chikungunya virus (Alphavirus family, Togaviridae Genus). It was initially identified in Tanzania in 1952 [1]. It is thought that Chikungunya outbreaks likely happened before the virus was identified as there are variable presentations of epidemic fevers with remarkable arthralgia. In the last 50 years, Chikungunya has re-emerged in several occasions in both Africa and Asia [2]. Aedes aegypti and Aedes albopictus mosquitoes are responsible for transmission of both Chikungunya and dengue [2]]and in Asia, have been identified as the primary vector in most urban dengue epidemics [3].

Chikungunya outbreaks were first noticed in 2008 in the northwest part of Bangladesh, affecting 39 people[4] followed by 196 cases in Nababgonj and Dohar subdistricts of Dhaka in 2011; additionally, sporadic cases were reported in 2013, 2014, and 2015, leading to a significant outbreak in December 2016[5]

The most severe outbreak of chikungunya in Bangladesh occurred between April and September 2017, outranking previous incidents, with over 13,000 clinically confirmed patients were reported in Dhaka alone and overall cases documented across 23 districts nationwide.[6]

In the year 2025, Chikungunya outbreak showing an enormous outbreak in Chattogram and a previous study done by Anamul et al.[7] rightly predicted about the outbreak in the year 2025. Concomitant Dengue and Chikungunya infection with few cases of Zika viral infection are prevailing during monsoon and post monsoon season. But there are rarity of documentation of Chikungunya in context of Chattogong. So the objectives of the present study is to document the clinical, serological and molecular data of Chikungunya cases in our context.

## Methods

This cross sectional observational study was conducted in the inpatient and out patient department of a tertiary care hospital of Chattogram, Bangladesh during a period of three months in the year 2025. A total of 120 clinically, serologically or by molecular assay positive cases were included in the study after informed written consent. Patients with prior diagnosis of other viruses were excluded. Co infection with or concomitant infection with Chikungunya and other viruses were included. Among all clinically suspected Chikungunya cases RT PCR for Chikungunya was done when patients come within the first 5-6 days of onset of fever. Again serological diagnosis by IgM or IgG were done if patients come after 6 days of fever or onwards. Clinical features were included as per previous study findings[6] and all common and uncommon clinical features were noted. Leucopenia was defined if WBC count is less then 4000/cumm, neutropenia was defined as it is <40% and lymphopenia was defined as it is <20%, >40 mm in 1^st^ hour was regarded as high ESR. After collection of all data it was compiled and analyzed by SPSS 25. Ethical clearance was taken from the Institutional Review Board(IRB) of Chattogram Maa O Shishu Hospital Medical College.

## Results

A total of 120 cases were studied in the present study.

**Table 1:** Gender distribution(N=120)

|  | Frequency | Percent |
| --- | --- | --- |
| Female | 63 | 52.5 |
| Male | 57 | 47.5 |
| Total | 120 | 100.0 |

Among the 120 patients female was more then the male (52.5% vs 47.5%)

**Table 2:**
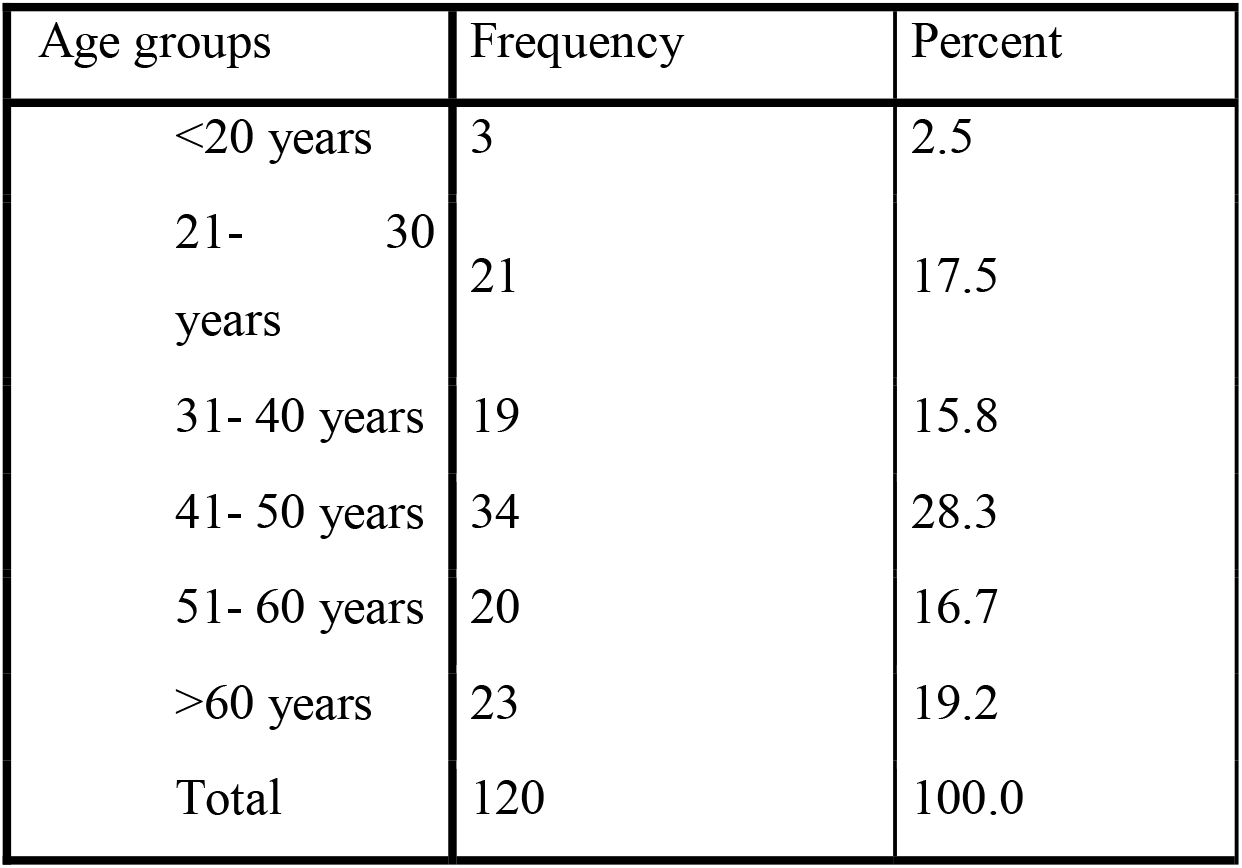
Age group distribution(N=120)

| Age groups | Frequency | Percent |
| --- | --- | --- |
| <20 years | 3 | 2.5 |
| 21- 30<br>years | 21 | 17.5 |
| 31- 40 years | 19 | 15.8 |
| 41- 50 years | 34 | 28.3 |
| 51- 60 years | 20 | 16.7 |
| >60 years | 23 | 19.2 |
| Total | 120 | 100.0 |

Table 2 showing age group distribution where mean age of the patients were found 45.34± 15. 37 years. Age group distribution were found even from 20 to 60 years or more. Among all 28.3%(34) patients were at age group 41 50 years which was highest.

**Table 3:** Clinical presentations(N=120)

| Features | Frequency | Percent |
| --- | --- | --- |
| Fever | 98 | 81.66 |
| Previous H/O fever or afebrile | 22 | 18.33 |
| Multiple joint pain | 88 | 73.33 |
| Ankle swelling and pain | 43 | 35.83 |
| Body aches | 52 | 43.33 |
| Rash | 41 | 34.16 |
| Nausea/Vomiting | 21 | 17.5 |
| Urticaria like skin lesion | 11 | 9.16 |
| Nose pigmentation | 5 | 4.16 |
| Others | 10 | 8.33 |

Table 3 showing different clinical presentations of chikungunya fever where some most common presentations were fever(81.66%), multiple joint pain (73.33%), ankle swelling and pain (35.83%), bodyaches (43.33%) and rashes (34.16%). Skin rash, nausea/vomiting, urticaria like skin lesions, nose pigmentation, ulcers, lymph node welling and painful morning stiffness were found as uncommon findings.

**Table 4:** Hematological findings(N=120)

| Features | Frequency | Percent |
| --- | --- | --- |
| Normal Hb | 114 | 95.00 |
| Raised ESR(>40mm in 1<sup>st</sup> hour) | 25 | 20.33 |
| Leucopenia | 22 | 18.33 |
| Neutropenia | 13 | 10.83 |
| Lymphopenia | 59 | 57.50 |
| Normal range platelet count | 115 | 95.83 |

Table 4 showing Hb% were found normal in 95% patients and high ESR was found in 25(20.33%) cases, leucopenia, lymphopenia and neutropenia was found in 18.33%, 57.50% and 10.83% respectively. Among all 115(95.83%) had normal ranged platelet.

**Table 5:** Serological and molecular findings(N-120)

| Findings | Frequency | Percent |
| --- | --- | --- |
| Chikungunya IgM +Ve | 66 | 55.00 |
| Chikungunya IgG +Ve | 7 | 5.83 |
| RT PCR for Chikungunya Positive | 42 | 35.0 |
| RT PCR for Chik/Dengue Positive | 5 | 4.16 |
| Influenza A + Ve | 1 | 0.8 |
| Influenza B + Ve | 2 | 1.6 |
| NS1 for dengue +ve | 6 | 5.00 |

\*Multiple response

Table 5 showing serological findings Chikungunya Ig M and IgG was found positive among 55.0% and 5.83% respectively. Among all 6(5%) case was found NS1 positive and three were found Influenza A or B positive as coinfection with Chikungunya. Regarding molecular data 42(35.0%) were found RT PCR for chikungunya positive and 5 cases was found for both Chikungunya and Dengue positive.

**Table 6:**
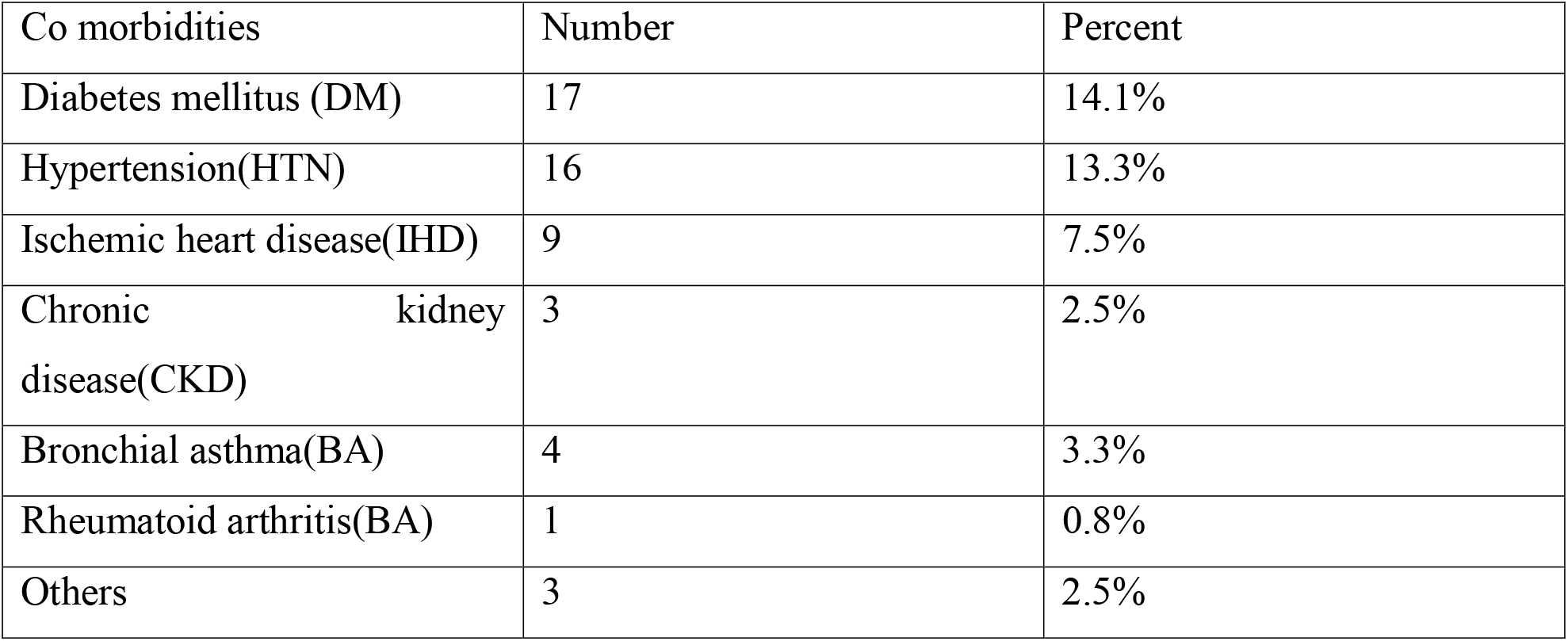
Comorbidities among the study patients(N=120)

| Co morbidities | Number | Percent |
| --- | --- | --- |
| Diabetes mellitus (DM) | 17 | 14.1% |
| Hypertension(HTN) | 16 | 13.3% |
| Ischemic heart disease(IHD) | 9 | 7.5% |
| Chronic kidney disease(CKD) | 3 | 2.5% |
| Bronchial asthma(BA) | 4 | 3.3% |
| Rheumatoid arthritis(BA) | 1 | 0.8% |
| Others | 3 | 2.5% |

Table showing among the study patients DM, HTN, IHD were found common comorbidities which were 14.1%, 13.3% and 7.5% respectively. One patient was found with RA which made dilemma with post chikungunya arthritis.

## Discussion

Chikungunya disease is characterized by fever and joint pain illness, but later can also lead to chronic arthritis and morbidity[1]. Both gender and all age group people are vulnerable to chikungunya fever as found in the present study where female were affected more then male 52.5% vs 47.5%. Young adults and older age groups all are vulnerable to infection. In a cohort of 55 individuals diagnosed with CHIKV infection in the year 2024[7] consisted of a disproportionate ratio of males to females (1.6:1), and a significant number of participants were predominantly between 18 and 30 years of age (n =19).

In the study most common presentations were found fever(81.66%) then multiple joint pain (73.33%), ankle swelling and pain (35.83%), bodyaches (43.33%) and rashes (34.16%). In a study done in Dhaka Bangladesh found most participants experienced fever onset within 2 days (n =34), indicating a rapid progression of symptoms after infection.. Next to fever most commonly reported symptoms included bone and joint pain, which was observed in a majority of the cases (n =51), followed by myalgia (n =48), headaches (n =45), and nausea and vomiting (n =35). It was found in that study[7] one participant, experienced gum bleeding, whereas another suffered from, lymph node swelling. In the present study skin rash, nausea/vomiting, urticaria like skin lesions, nose pigmentations ulcers, lymph node welling morning stiffness were found as uncommon findings. Generally, chikungunya is a self-limiting viral infection with fever, arthritis and rash. Some patients may have long duration joint symptoms [8] that requered different rheumatological investigations to exclude SPA or RA. Besides these, Chikungunya can involve various body systems, including central nervous system and cardiovascular system and rarely may be fatal one[9]. Also it was also found non fatal in the present study. Again we found no neurological and cardiovascular symptoms among all patients. Comorbidities were not found to influence in the progression and severity of symptoms in the present study

During the present outbreak in this year 2025 at Chittagong, Bangladesh, clinical and epidemiological criteria were used to make a probable diagnosis of chikungunya fever, but serological and molecular confirmation were used for the fulfillment of laboratory criteria irrespective of clinical presentation. RT-PCR was done in first week and IgM/IgG appeared after day 10 of symptom onset and which lasted for weeks to months. These serological and molecular findings were also described in a study done in 2015[10]. In the present study we found Raised ESR and presence of lymphopenia are important in laboratory differentiation from dengue found in our study. If patient comes early, Dengue NS 1 test may be done to exclude dengue in our study some patients found positive of it concomitantly. Interestingly few patients were also found flu virus positive.

In our study only RT PCR for Chikungunya and Dengue were done and sequencing was not done due to unavailability of resources. In a study done in India[11] phylogenetic analysis revealed that in the strains they analyzed was East Central South African (ECSA) genotype. Emergence of a variant strain was observed in the year 2016, which became the predominant strain in this region in 2017. The strains showed significant identity with recent New Delhi strains of 2015 and 2016 and Bangladesh strains of 2017. Also in a study done in 2024[7] among Dhaka Bangladesh samples where genetic analysis of CHIKV strains revealed an evolutionary shift, with the 2017 strain being genetically distinct from earlier outbreaks, suggesting an adaptive response that may have contributed to its widespread presence[12]This pattern suggests that the resurgence of CHIKV cases in late 2024 could signify the early stages of another substantial outbreak in 2025.

Sequencing and phylogenetic study of Chikungunya could not be done and also long term followup findings was not included which were some limitations of the present study.

In conclusion, chikungunya is responsible for significant morbidities with fever and arthralgia/arthritis with rashes. A multicenter study with long term followup findings in a bigger representative samples are needed to get a true picture of present reemerging Chikungunya fever in the context of Bangladesh.

## Data Availability

Data are available on Request

